# Multimodal Wearable System for Objective Assessment of Dynamic Rotational Knee Biomechanics Following ACL Injury and Reconstruction: A Clinical Validation Study Using Ensemble Deep Learning

**DOI:** 10.64898/2026.05.08.26352706

**Authors:** Jiya Dutta, David Tan Yuan Yu, Lai Kah Weng, Jeffrey Zhu, Chia Zi Yang

**Affiliations:** PRECIX Pte Ltd (Precix.io), 2 College Road, Singapore 169850, Singapore; Diagnostics Development Hub (DxD Hub), Agency for Science, Technology and Research (A^*^STAR), 3 Biopolis Drive, Singapore 138627, Singapore; College of Computing, Data Science, and Society, University of California, Berkeley, Warren Hall, 2195 Hearst Avenue, Berkeley, CA 94720-1786, USA; Department of Orthopaedic Surgery, Singapore General Hospital (SGH), Outram Road,Singapore 169608, Singapore

## Abstract

**Background:** The clinical assessment of knee stability after an Anterior Cruciate Ligament (ACL) injury is routinely conducted via operator-dependent physical examination tests (i.e. pivot shift) and standardized patient-reported outcomes. Unfortunately, both are unable to perceive and quantify the subtle rotational biomechanical deficiencies from an ACL tear. Although specialized laboratory-based motion capture systems may provide objective measurements, they are found in research institutions and thus, are not suitable for clinical use. In contrast, GATOR PRO is a clinic-based multimodal wearable sensor system that uses a machine learning (ML) model (ensemble deep learning) to differentiate and classify its data outputs for assessing in-vivo dynamic rotational knee stability.

**Objective:** The purpose of this study is to validate the deep machine learning model and its performance used in GATOR PRO, which integrates knee-mounted Inertial Measurement Units (IMUs) with ultrasound images to derive high-fidelity in-vivo biomechanical rotational data. Based on this data collected by the GATOR PRO, it is hypothesized that the model can effectively classify knee stability after ACL injury and reconstruction.

**Methods:** This prospective clinical study at Singapore General Hospital (SGH) (CIRB 2019/2766, PDPA-compliant) aimed to enroll 60 patients (30 ACL-deficient, 30 ACL-reconstructed ≥6 months post-surgery). At the halfway point of the clinical trial, 29 patients (8 ACL-deficient, 21 ACL-reconstructed ≥6 months post-surgery) were recruited through physician referral at SGH outpatient clinics to perform standardized chair-stand tests. An ensemble deep learning model that combines convolutional (EfficientNet) and time-series (InceptionTime) classifiers is used to output binary stability classifications (ACL-deficient/ACL-reconstructed). The model’s performance was evaluated using 10-fold stratified cross-validation with patient-wise splitting, repeated across 100 random seeds to assess variability.

**Results:** At the halfway point of the trial, the ensemble model performance with regard to the Receiver Operating Characteristic area under the curve (ROC-AUC) was 0.8365 (SD: 0.042, p-value < 0.001), and the classification accuracy was 75.9% (SD: 3.2%) when the model was tested on the 29 CIRB-approved patients. For the ACL-reconstructed class, the performance indicators were as follows: precision 71.4%, recall 93.8%, F1-score 81.1%. For the ACL-deficient class, the indicators were: precision 87.5%, recall 53.8%, F1-score 66.7%.Against the clinical pivot shift test’s low sensitivity (24-32%), the model delivers an almost 2X better sensitivity (53.8%)[2, 3], with a comparable specificity (93.8% vs. 90-98%)

**Conclusion:** The multimodal machine learning model was able to perform at a level that was relevant to clinical classification (AUC-ROC 0.8365, accuracy 75.9%) in differentiating between ACL-deficient and ACL-reconstructed knees. Moreover, the model demonstrated far superior sensitivity than previously published estimates for manual pivot shift testing (53.8% vs. 24-32%). These findings demonstrate that rotational knee instability can be reliably differentiated in clinical settings with a ML model deployed on GATOR PRO data.

## INTRODUCTION

### Clinical Background and Motivation

The anterior cruciate ligament (ACL) is the primary stabilizer against anterior tibial translation and rotational movements during dynamic activities such as pivoting.. Injuries to the ACL are among the most frequent and severely disabling knee problems, especially in the active population, with 200,000 cases yearly in the United States alone [4, 5]. In addition to the loss of function, ACL deficiency causes changes in the biomechanics of the joint, which in turn increase the risk of the secondary damage to the meniscus and cartilage, leading to early post-traumatic osteoarthritis in up to 50% of the patients within 10-15 years of the initial trauma [6, 7].

Although ACL injury is of major clinical concern, current assessment methods remain inadequate for quantifying subtle rotational deficiencies which are important for prognosticating eventual outcomes. The use of standard clinical tools involving the manual pivot shift and Lachman tests is subjective by nature and shows a substantial variability between examiners [2]. At the same time, patient questionnaires like the International Knee Documentation Committee (IKDC) score, although they are able to reflect the functional deficit, cannot objectively measure the biomechanical ones. Most importantly, these traditional approaches are insensitive to measuring rotational instability, the primary functional deficit after ACL injury, which manifests during dynamic motion rather than static examination.

Precise biomechanical data can also be obtained by laboratory-based motion capture systems that make use of optical tracking and force plates but these systems have not been widely adopted in clinical practice yet. These research tools require dedicated space, extensive preparation, specialized personnel, and analysis times ranging from hours to days.Therefore, these systems are considered to be inappropriate for clinical practice at places like SGH where fast and point-of-care assessment is a must.

Subjective methods predominate in the evaluation of ACL and this has resulted in several serious drawbacks. According to systematic reviews [2, 3], the pivot shift test, which is the gold standard in the evaluation of rotational instability, is characterized by sensitivity varying from 24 to 32% only in patients in clinical settings, with specificity of 90-98%. The high false-negative rate results in a significant number of patients with functional instability remaining undetected during clinical examination. Also, the quality of the test depends mostly on the experience of the examiner, the level of relaxation of the patient, and the presence or absence of muscle guarding, thus leading to variations in the results even when performed by skilled orthopedic surgeons.

The machine learning approach presented here addresses these limitations through integration of multimodal sensing and deep learning classification. Through measurements on cadavers (establishing measurement accuracy), experiments on living subjects (verifying real-world precision), and usability testing (ensuring clinical workflow compatibility), GATOR PRO [1], the wearable sensor system that provides input data to the model, was brought to the point of readiness. This study, conducted at SGH with the analytical support from UC Berkeley and National University of Singapore, serves as evidence of the performance of the machine learning model in detecting the stability of the knee and also compares its diagnostic accuracy with that of the clinical benchmarks, including the pivot shift test.

## METHODS

### Study Design and Ethics Approval

This clinical validation study was conducted at the Department of Orthopaedic Surgery, Singapore General Hospital. Approval from the Centralised Institutional Review Board (CIRB 2019/2766) was granted in accordance with Singapore’s Personal Data Protection Act (PDPA) regulations. The study aimed to evaluate the system’s ability to objectively detect changes in range of motion and rotation of the femur relative to tibia in ACL-deficient and ACL-reconstructed knees [1]. At the time of the analysis (29 out of 60 planned patients enrolled), the study protocol involved the use of wearable sensors to collect data onsite, which was then uploaded to a secure cloud platform for AI analysis. Thus, no additional computing infrastructure was required at the clinical site. All the individuals who took part in the study signed the informed consent form with SGH’s Clinical Research Coordinators (CRCs) before being enrolled, having been fully informed about the use of data, privacy safeguards, and the terms of voluntary participation.

### Data Collection Protocol

To address the limitations of Soft Tissue Artifacts (STA) in kinematic measurements [8, 9, 10], an integrated multi-modal wearable sensor system was utilized. This system combined inertial sensing with real-time ultrasonography to capture high-fidelity skeletal motion.

#### Inertial Measurement Units (IMUs)

IMUs each consisting of 3-axis accelerometers and 3-axis gyroscopes were used to capture motion at 100Hz during dynamic tasks [1].

#### Ultrasound Transducers

To mitigate errors introduced by skin motion [9, 10], an integrated ultrasound-based tracking system was employed to track the underlying skeletal motion directly [1]. The workflow for artifact correction followed a computational sequence:

1. Acoustic Visualization and Segmentation: The femoral cortex was visualized in real-time. A proprietary bone segmentation mask was applied to the acoustic images to isolate the hyperechoic skeletal boundary from the adjacent soft tissue layers.
2. Dynamic Bone Tracking: The system utilized skeletal landmarks and contour features to track the precise temporal displacement of the bone by cross-referencing subsequent image frames.
3. Algorithmic Calibration: The captured bone displacement was synchronized with IMU data and processed via a proprietary calibration algorithm. This algorithm was previously validated through computational simulations, cadaveric research [15], and robotic bench testing [1].

Procedural Standardization: The sensors were applied to both legs by trained members of the research team following a standardized positioning protocol to ensure anatomical consistency across all subjects.

### Functional Assessment Protocol

This functional assessment was preferred to the traditional clinical examination maneuvers (e.g., pivot shift) because it:

- Is a natural, dynamic movement directly related to the activities of daily living
- Can be done consistently by patients with different functional levels
- Neuromuscular control mechanisms, which are the main compensators for ligamentous deficiency, get activated
- No specialized equipment is needed

Patients during testing were asked to perform 3-5 times chair-stand movement while the sensor system was continuously recording bilateral IMU signals and ultrasound images. This repetition scheme offered several data samples per patient while lessening the fatigue effects.

## MACHINE LEARNING MODEL DEVELOPMENT AND TECHNICAL ARCHITECTURE

The classification system employs a deep learning architecture, using the most complementary data that comes from multimodal sensor inputs.

### Component Model 1: EfficientNet-Based Convolutional Neural Network

- Architecture: EfficientNetB0 backbone pretrained on ImageNet. Incorporates Long Short-Term Memory (LSTM) network for temporal sequence modeling.
- Input: Ultrasound images after being preprocessed (224×224 pixels, grayscale)
- Training Status: 10 models were trained (1 for each random seed, seeds 0-9) using transfer learning with a maximum of 100 epochs and early stopping based on the validation AUC-ROC convergence.

To address the limited dataset, a three-stage transfer learning process was implemented.

1. Epoch 1-15: Backbone with LSTM (LR: 1 x 10^-3^)
2. Epoch 16-40: Final 2 EfficientNet blocks (LR: 1 x 10^-4^)
3. Epoch 41-100: Full fine-tuning (LR: 1 x 10^-5^)

Generalisation was supported by stochastic augmentation intensity (brightness, contrast etc.) and temporal (frame dropout). Regularisation included dropout (0.3-0.4), AdamW Optimiser with weight decay (0.05) and label smoothing (0.1). To handle small-sample variance, an early stopping criterion was applied, such that training was halted if the 5-epoch exponential moving average of the validation AUC-ROC failed to improve for 20 epochs. A minimum of 30 epochs was enforced to ensure the completion of the first transfer learning stage. The model weights yielding the highest smoothed validation AUC were restored for testing.

### Component Model 2: InceptionTime-Based Time-Series Classifier

- Architecture: The architecture is based on the InceptionTime framework (Fawaz et al., 2020), which is implemented in the open-source tsai library.
- Input: 12-channel IMU time series resampled to 1000 timesteps per sequence.
- Training Status: 100 models were trained (10 patient-wise data splits x 10 random seeds per split) for 150 epochs each.
- InceptionTime was designed for time-series classification and has achieved state-of-the-art performance on benchmark datasets.

### Ensemble Framework Integration

The final classification system takes advantage of the different strengths of each sensing method:

1. Ultrasound allows for correction of soft-tissue artifacts [1], whereas IMU sensors provide temporal kinematics through functional movements.
2. The ensemble integration uses probability-level fusion:

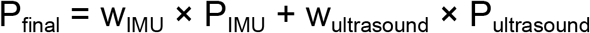

where:
  - P_IMU_ is the average probability of an ACL-deficient knee over 100 InceptionTime models
  - P_ultrasound_ is the average probability of an ACL-deficient knee over 10 EfficientNet models Current Configuration: w_IMU_ = 0.70; w_ultrasound_ = 0.30
  - The InceptionTime component was fully trained (100 models, 150 epochs each) while the EfficientNet component still needs extended training for full optimization. The weighting thus gives more weight to the validated IMU predictions and at the same time incorporates ultrasound.
  - Both the InceptionTime and EfficientNet components were trained using predefined maximum epoch limits with early stopping based on validation loss. The ensemble weighting reflects the stronger standalone performance of the IMU-based model observed in this study.
  - The weighting strategy prioritises the IMU modality while incorporating complementary information from the ultrasound component to improve robustness against soft tissue artifacts.
  - The ensemble model outputs binary classification scores with confidence measures as a degree.

Binary classification uses a threshold of 0.5 on the ACL-deficient probability:

P_final > 0.5 → ACL-deficient

P_final ≤ 0.5 → ACL-reconstructed

A confidence score is given and can be high (>0.73), moderate (0.5-0.73), or low (0.32-0.5).

## RESULTS

### Classification Performance

At the trial midpoint, 29 patients had been enrolled (8 ACL-deficient, 21 ACL-reconstructed ≥6 months post-surgery). The ensemble model achieved the following knee stability classification performance:

### Comparison with Clinical Benchmarks

The performance of the ensemble model was compared to that of the manual pivot shift test, which is the clinical gold standard for assessing rotational knee instability [2, 3]:

#### Performance Comparison

- Sensitivity: 53.8% (ensemble model) vs. 32% (pivot shift test [2] from systematic reviews)
- 68% relative improvement in identification of unstable knees
- Specificity: 93.8% (ensemble model) vs. 90-98% (pivot shift test [2, 3])
- Comparable specificity while substantially improving sensitivity.The high specificity shows that the false positive rate is still at an acceptable level.
- Implications:
  - Improved identification of at-risk patients
  - Objective assessment when the clinical examination is equivocal or limited by factors of the patient (pain, guarding, anxiety)

## DISCUSSION

### Principal Findings and Clinical Significance

This paper reports the evaluation of a multimodal wearable sensor system [1] for objectively classifying knee stability post ACL injury and reconstruction. The deep learning ensemble framework was able to achieve clinically relevant discriminative performance (AUC-ROC 0.8365, accuracy 75.9%) in a pilot cohort of 29 patients

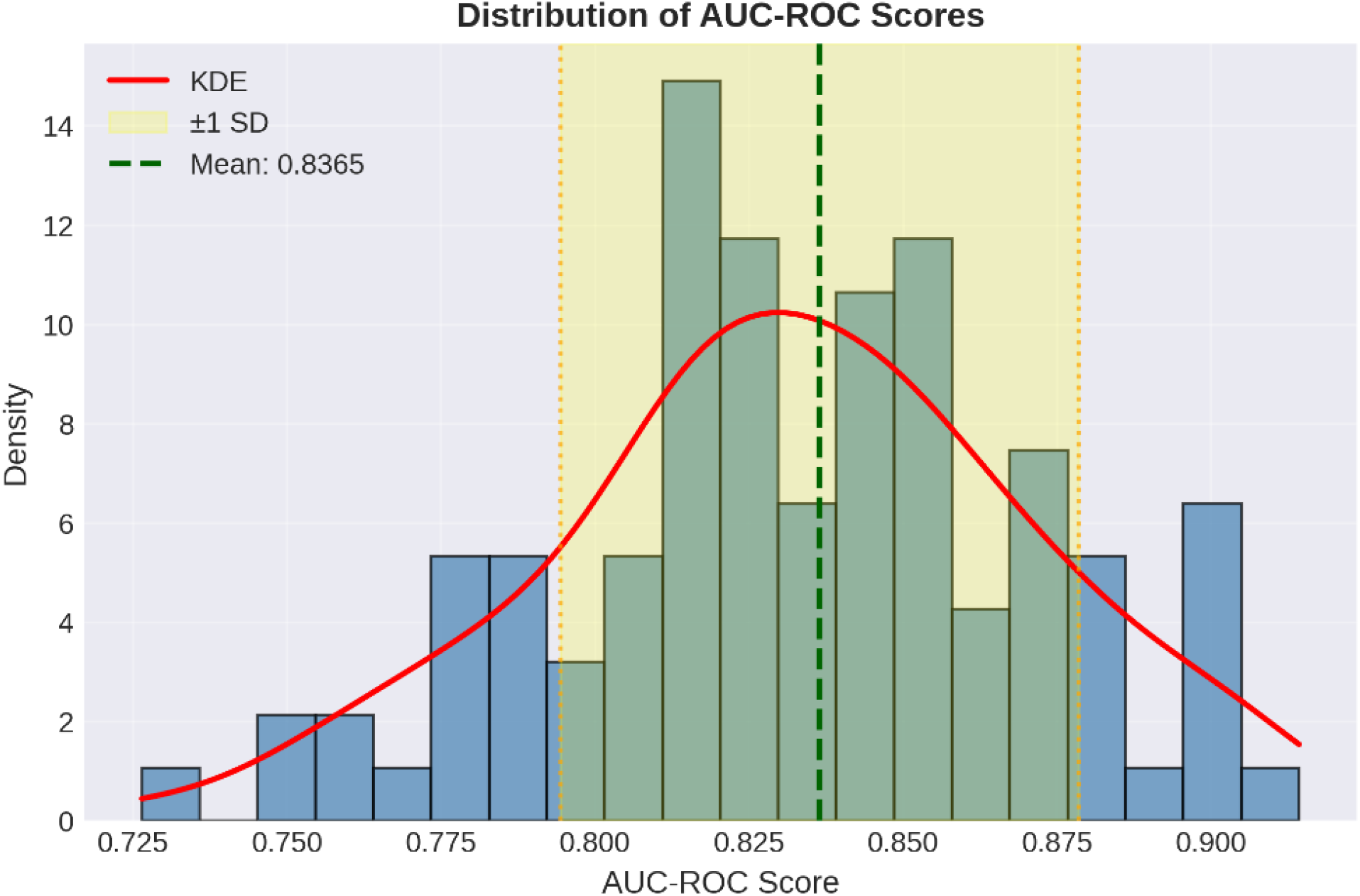

### Key Performance Achievements

The achieved sensitivity of 53.8% is a 68% relative improvement over manual pivot shift testing (32% sensitivity [2] from systematic reviews) at the same time a similar specificity (93.8% vs. 90-98% [2, 3]) is maintained. This performance profile addresses a key limitation of the ACL assessment presence of rotational instability, potentially enabling identification of patients who might be missed by conventional clinical examination. The ROC-AUC of 0.8365 (p-value < 0.001) signals that the model has a good capacity for discrimination, and this level is considered clinically useful for medical decision support systems. The classification accuracy of 75.9%, which is stable even under repeatedcross-validation, is evidence of strong generalization capability. These results represent a pilot feasibility analysis designed to evaluate model discriminative capacity and ensemble performance, prior to a large-scale clinical validation.

The multimodal method tackles the major problems of the previous wearable systems that have been identified.Ultrasound imaging provides skeletal reference frames to correct for soft tissue artifacts [1, 9, 10], which is the main source of error in skin-mounted IMU measurements. Functional assessment through chair-stand tests records the dynamic knee behavior during weight-bearing transitions that are typical of daily living activities. Moreover, the system offers standardized measurements that make it possible to compare reliably different time points and clinical sites, in contrast to examiner-dependent manual tests.

### Clinical Applications

The objective assessment may be of use in clinical practice.

1. Quantitative stability metrics can be a useful pre-operative tool to guide surgeons in customizing the ACL surgery for their patients, e.g. including a lateral extra-articular procedure.
2. Monitoring during rehabilitation provides objective functional recovery data, thus enabling the clinician to decide the return-to-sport timeframe and identify the patients with biomechanical deficits.
3. Quantitative assessment can reveal patients with persistent instability who might not be apparent on clinical examination and, thus, constitute a high-risk group for secondary joint damage [6,7].
4. Use of standardized objective measurement allows for a more rigorous evaluation of different ACL reconstruction techniques, rehabilitation protocols, and surgical decisions.
5. Limitations of Analysis Multimodal Integration Status: The validation has been completed for the IMU-based classification using the current system. To reach the performance level of the InceptionTime (IMU) component, the EfficientNet (ultrasound) part needs more training:
  a. InceptionTime: 100 fully trained models (10 data splits × 10 training seeds, 150 epochs each)
  b. EfficientNet: 10 models trained (10 epochs each), requiring extension to 100-150 epochs The following research will not only finish EfficientNet training but also systematically optimize weight through ablation studies.
  This shortcoming does not lessen the clinical value of IMU-based assessment which achieves robust discrimination (AUC 0.8365) beyond manual examination benchmarks [2, 3].

## CONCLUSIONS

The study evaluated ensemble deep learning applied to wearable sensor data for knee stability classification after ACL injury and reconstruction. At the trial midpoint (29 patients), the system demonstrated performance that could be considered clinically meaningful: ROC-AUC of 0.8365, classification accuracy of 75.9%, and sensitivity relative improvement of 68% over manual pivot shift testing (53.8% vs. 32%) while specificity was comparable (93.8%). There are current limitations such as incomplete enrollment (48% of target), class imbalance (27.6% ACL-deficient), single-site validation, and binary classification framework. To confirm clinical utility, further recruitment towards a balanced 60-patient enrollment, multi-site validation, and longitudinal outcome tracking are necessary. These results are a step towards the use of wearable sensor-based methods and machine learning to objectively quantify dynamic knee biomechanics and thus have the potential to be a good supplement for manual clinical examination.

## Data Availability

All data produced in the present study are available upon reasonable request to the authors.

## Funding

This study was supported by the Diagnostics Development Hub (DxD Hub), Agency for Science, Technology and Research (A*STAR), Singapore, through a collaborative research agreement with PRECIX Pte Ltd and Singapore General Hospital (SGH Ref: RA2020-035). DxD Hub’s contribution was funded in part by the Singapore National Research Foundation (NRF) innovation cluster programme. PRECIX Pte Ltd provided the GATOR devices and associated platform for the clinical study at its own cost. SGH provided clinical infrastructure, patient recruitment, and study team support. The funders participated in study design and project management.

## Conflict of Interest

Jiya Dutta is an employee of PRECIX Pte Ltd, the company that developed and provided the GATOR device used in this study. Lai Kah Weng is an employee of PRECIX Pte Ltd, the company that developed and provided the GATOR device used in this study. David Tan Yuan Yu is an employee of the Diagnostics Development Hub (DxD Hub), A*STAR, which provided funding and project management support for this study under a collaborative research agreement with PRECIX Pte Ltd and Singapore General Hospital. Chia Zi Yang is a clinician at the Department of Orthopaedic Surgery, Singapore General Hospital, which participated in this study as an institutional co-investigator under a collaborative research agreement with PRECIX Pte Ltd and DxD Hub, A*STAR. Jeffrey Zhu declares no conflicts ofß interest.

## Acknowledgments

We thank PRECIX Pte Ltd, A*STAR’s Diagnostics Development Hub, SGH’s Department of Orthopaedic Surgery, and the Department of Bioengineering at UC Berkeley for their collaborative efforts in developing and validating GATOR PRO. Special thanks to:

## Clinical Team

- Dr. Deborah Huang and Dr. Lee Kong Hwee (SGH) for patient referrals and clinical expertise
- William Yeo from Orthopaedic Diagnostic Center (SGH) for overall patient enrollment coordination
- Shameemah Binte Mohamed Fouze and Adam Farid Tang Ming Tang (Clinical Research Coordinator team, SGH) for driving patient recruitment and obtaining informed consent

## Technical Team

- Justin Wong (BSc, School of Computing, National University of Singapore) and Xu Yunhe (BSc, Faculty of Science, National University of Singapore) for contributions to model development and validation analysis

## Leadership

- Lim Qing Ru, CEO of PRECIX Pte Ltd, for project leadership and strategic direction

## Participants

- All study participants who generously contributed their time and data to advance ACL assessment research
- Clinical staff at SGH for supporting trial implementation

## APPENDIX

Patient Demographics

| Characteristic | ACL-Deficient (n=8) | ACL-Reconstructed (n=21) | Total (n=29) |
| --- | --- | --- | --- |
| Age (years) |  |  |  |
| Range | 18-65 |  |  |
| Time Since Injury/Surgery |  |  |  |
| ACL-deficient (months) | Variable | N/A | - |
| Post-reconstruction (months) | N/A | ≥6 | - |
| Data Quality |  |  |  |
| Sessions with ≥80% signal quality | 8 (100%) | 21 (100%) | 29 (100%) |
| Complete IMU sequences | 8 (100%) | 21 (100%) | 29 (100%) |
| Valid ultrasound images | 8 (100%) | 21 (100%) | 29 (100%) |

Model Architecture

| Component | InceptionTime (IMU) | EfficientNet (Ultrasound) |
| --- | --- | --- |
| BASE ARCHITECTURE |  |  |
| Framework | tsai library | PyTorch + Lightning |
| Base Model | InceptionTime | EfficientNet-B0 + LSTM |
| Input Modality | 12-channel IMU time series | Ultrasound image sequences |
| Input Dimensions | (B, 12, 1000) | (B, 50, 1, 224, 224) |
| Feature Extraction | Multi-scale temporal convolutions | Spatial (CNN) + temporal (LSTM) |
| Output Dimension | 2 classes<br>(ACL-deficient/ACL-reconstructed) | 2 classes<br>(ACL-deficient/ACL-reconstructed) |
| TRAINING CONFIGURATION |  |  |
| Number of Models | 100 | 10 |
| Data splits | 10 | 1 (per seed) |
| Training seeds per split | 10 | 10 |
| Training Epochs | 150 | 10* |
| Batch Size | 6 | 1 |
| Optimizer | Adam | AdamW |
| Learning Rate | 0.2 (max) | 0.001 |
| LR Schedule | fit_one_cycle (warmup + cosine) | Cosine with warmup |
| Weight Decay | Not specified | 0.05 |
| Loss Function | Cross-entropy | Cross-entropy |
| Class Weights | Not applied (stratified sampling) | Not applied (balanced sampling) |
| Checkpoint Selection | Min validation loss (after epoch 100) | Min validation loss (not implemented) |
| Regularization | Per-sample batch standardization | Dropout (p=0.2) |
| DATA PREPROCESSING |  |  |
| Sequence Length | 1000 timesteps | 50 frames |
| Normalization | Per-sample z-score (by_sample=True) | ImageNet statistics (mean/std per channel) |
| Augmentation | None | Color jitter (brightness, contrast) |
| Train/Validation Split | 80:20 (patient-wise) | 80:20 (patient-wise) |
| COMPUTATIONAL REQUIREMENTS |  |  |
| Training Time per Model | ~1-1.5 hours | ~30-45 minutes |
| Total Training Time | ~100-150 GPU hours | ~5-7.5 GPU hours |
| Hardware | CUDA GPU | CUDA GPU |
| Mixed Precision | Not specified | Enabled (FP16) |
| TRAINING STATUS |  |  |
| Completion Status | ✓ Fully trained | Requires extension |
| Validation Protocol | ✓ 100-run CV | Single split per seed |
| Ready for Deployment | ✓ Yes | Framework only |
| Contribution to Ensemble | 70% | 30% |

Performance Results

| Metric | Value (Mean ± SD) | 95% CI |
| --- | --- | --- |
| ROC-AUC | 0.8365 ± 0.042 (p-value < 0.001) | [0.828, 0.845] |
| Range across 100 runs | [0.72, 0.93] | - |
| Classification Accuracy | 75.9% ± 3.2% | [74.6%, 77.2%] |
| Macro-averaged F1-score | 73.90% | - |
| Weighted-averaged F1-score | 74.60% | - |

| Metric | ACL-Reconstructed ("Healthy")<br>n=21 patients | ACL-Deficient ("Unhealthy") n=8<br>patients |
| --- | --- | --- |
| CLASSIFICATION METRICS |  |  |
| Precision | 71.40% | 87.50% |
| Interpretation | When model predicts "healthy,"<br>correct 71.4% of time | When model predicts "unhealthy,"<br>correct 87.5% of time |
| Recall (Sensitivity) | 93.80% | 53.80% |
| Interpretation | Correctly identifies 93.8% of truly<br>reconstructed knees | Correctly identifies 53.8% of truly<br>deficient knees |
| Specificity | 53.80% | 93.80% |
| F1-Score (harmonic mean) | 81.10% | 66.70% |
| CONFUSION MATRIX |  |  |
|  | Actual ACL-Reconstructed<br>(n=21) | Actual ACL-Deficient (n=8) |
| Predicted ACL-Reconstructed | 20 (True Negative) | 4 (False Negative) |
| Predicted ACL-Deficient | 1 (False Positive) | 4 (True Negative) |

Comparison

| Assessment Method | Sensitivity (%) | Specificity (%) | AUC-ROC | Reference |
| --- | --- | --- | --- | --- |
| GATOR PRO MULTIMODAL ENSEMBLE |  |  |  |  |
| Current study | 53.8 | 93.8 | 0.837 | Present study |
| MANUAL CLINICAL EXAMINATION |  |  |  |  |
| Pivot Shift Test<br>(clinic setting) | 24-32 | 90-98 | - | Benjaminse 2006;<br>Lording 2019 |
| Lachman Test | 86 | 91 | - | Makhmalbaf 2013;<br>Reiman 2013 |
| Anterior Drawer<br>Test | 48 | 87 | - | van Eck 2013 |
| Combined Manual<br>Tests (Lachman +<br>Anterior Drawer) | Variable | Variable | - | Multiple sources |
| INSTRUMENTED<br>DEVICES |  |  |  |  |
| KT-1000<br>Arthrometer<br>(anterior<br>displacement) | 63-92 | 71-99 | - | Daniel 1985;<br>Panariello 1991 |
| Electromagnetic<br>System (pivot shift<br>quantification) | Variable | Variable | - | Hoshino 2012 |
| RELATIVE<br>PERFORMANCE |  |  |  |  |
| GATOR vs. Pivot<br>Shift |  |  |  |  |
| Sensitivity<br>improvement | 0.68 | - | - | Present study vs.<br>Benjaminse 2006 |
| (53.8% vs. 32%) |  |  |  |  |
| Specificity<br>comparison | Comparable | - | - | Present study vs.<br>literature |
| (93.8% vs. 90-98%) |  |  |  |  |

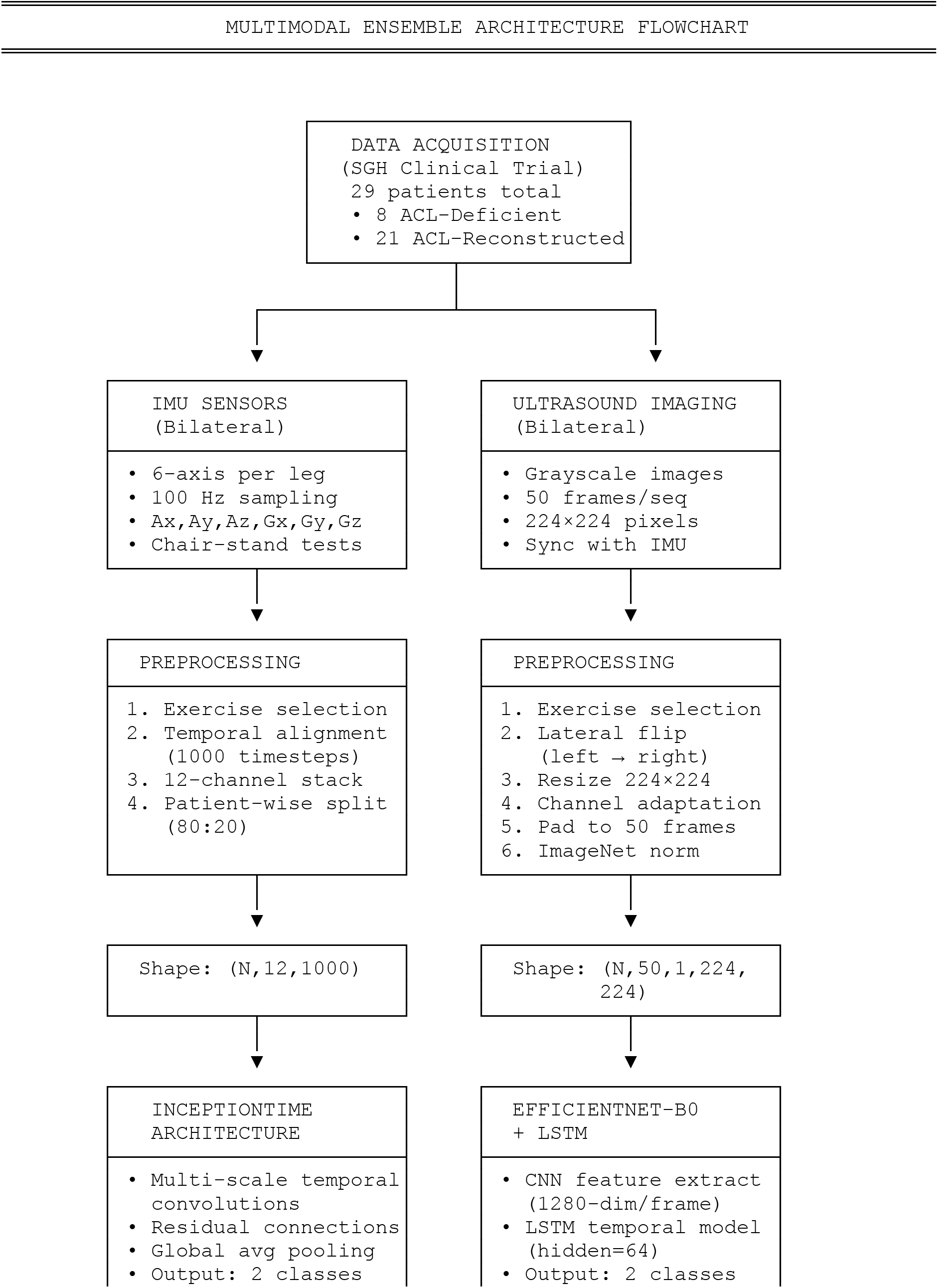
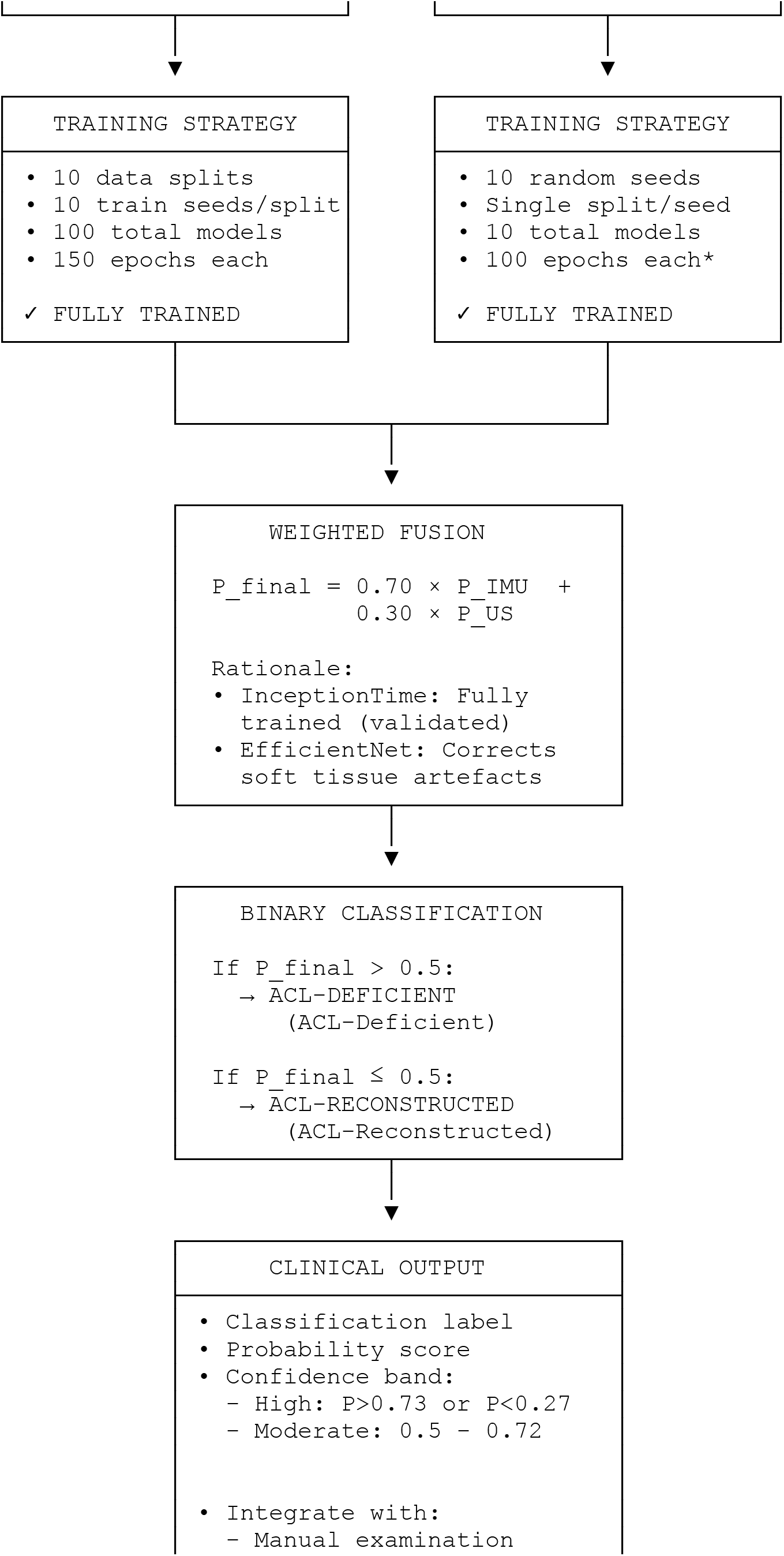
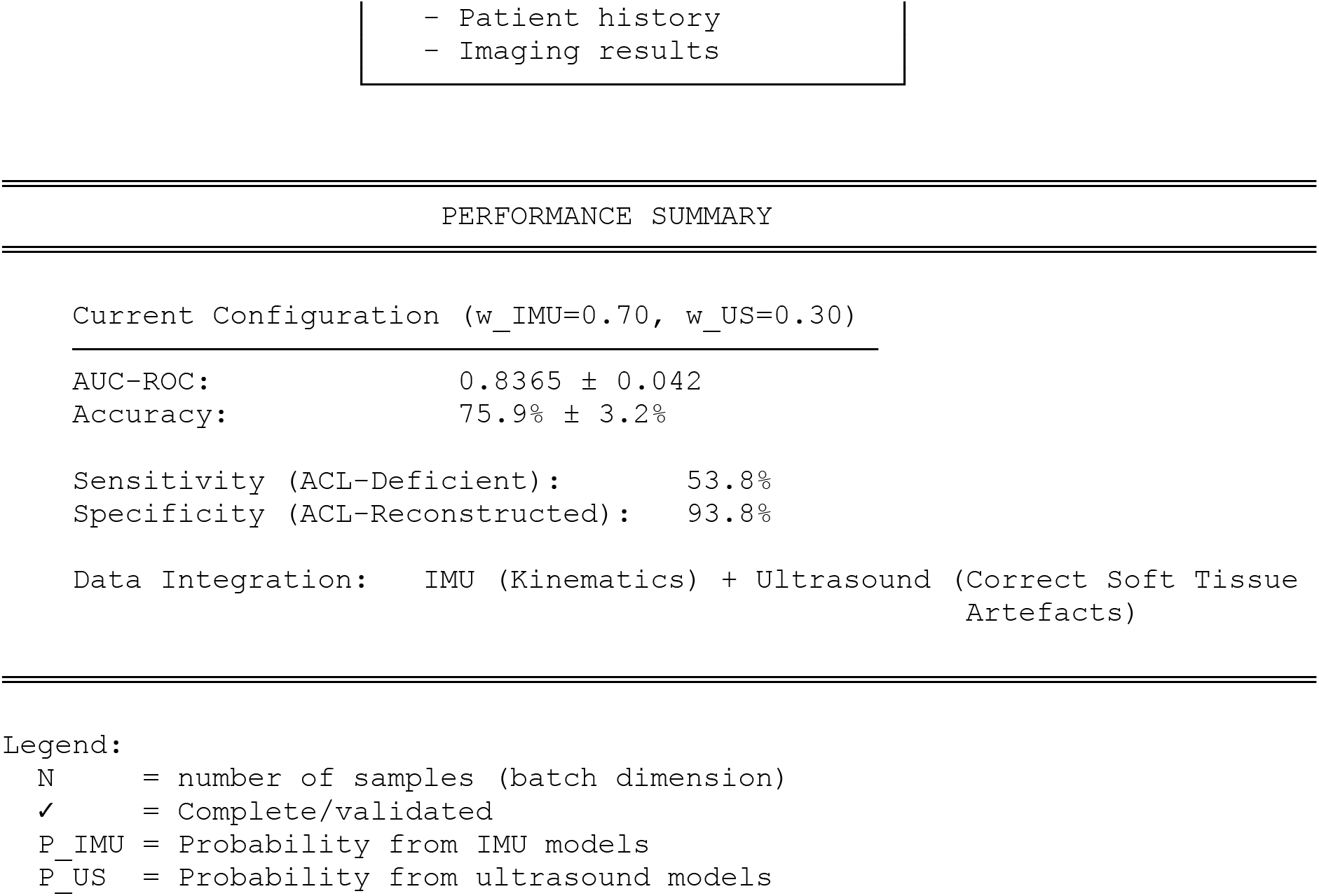

## Notes

### Author Declarations

CIRB 2019/2766 of Singapore General Hospital gave ethical approval for this work

